# Social deprivation and SARS-CoV-2 testing: a population-based analysis in a highly contrasted Southern France region

**DOI:** 10.1101/2023.02.09.23285721

**Authors:** Jordi Landier, Léa Bassez, Marc-Karim Bendiane, Pascal Chaud, Florian Franke, Steve Nauleau, Fabrice Danjou, Philippe Malfait, Stanislas Rebaudet, Jean Gaudart

## Abstract

**Background:** Testing was the cornerstone of the COVID-19 epidemic response in most countries until vaccination became available for the general population. Social inequalities generally affect access to healthcare and health behaviours, and COVID-19 was rapidly shown to impact deprived population more drastically. In support of the regional health agency in Provence-Alpes-Côte d’Azur (PACA) in South-Eastern France, we analysed the relationship between testing rate and socio-demographic characteristics of the population, to identify gaps in testing coverage and improve targeting of response strategies.

**Methods:** We conducted an ecological analysis of SARS-CoV-2/COVID-19 testing rate in the PACA region, based on data aggregated at the finest spatial resolution available in France (IRIS) and by periods defined by public health implemented measures and major epidemiological changes. Using general census data, population density, and specific deprivation indices, we used principal component analysis followed by hierarchical clustering to define profiles describing local socio-demographic characteristics. We analysed the association between these profiles and testing rates in a generalized additive multilevel model, adjusting for access to healthcare, presence of a retirement home, and the age profile of the population.

**Results:** We identified 6 socio-demographic profiles across the 2,306 analysed IRIS spatial units: privileged, remote, intermediate, downtown, deprived and very deprived (ordered by increasing social deprivation index). Profiles also ranged from rural (remote) to high density urban areas (downtown, very deprived). From July 2020 to December 2021, we analysed SARS-CoV-2/COVID-19 testing rate over 10 periods. Testing rates fluctuated strongly but were highest in privileged and downtown areas, and lowest in very deprived ones. The lowest adjusted testing rate ratios (aTRR) between privileged (reference) and other profiles occurred after implementation of a mandatory healthpass for many leisure activities in July 2021. Periods of contextual testing near Christmas displayed the largest aTRR, especially during the last periods of 2021 after the end of free convenience testing for unvaccinated individuals.

**Conclusions:** We characterized in-depth local heterogeneity and temporal trends in testing rates and identified areas and circumstances associated with low testing rates, which the regional health agency targeted specifically for the deployment of health mediation activities.

## Introduction

In recent years, social epidemiology has made a significant contribution to showing and explaining health inequalities. Some authors are now advocating an integrative approach based on interdisciplinarity and integrating social science theory more deeply [1]. The role played by socio-economic status (SES) in access to care, use of care and health behaviors [2] must thus be taken into account to study the disparities observed within the population in the recent COVID-19 pandemic, especially in access and use of diagnostic tests. From the first months of the pandemic, COVID-19 was rapidly shown to impact more dramatically populations who were already affected by socio-economic deprivation in European countries [3, 4].

During the 18 months after the first hard lockdown in France, mass testing, tracking and isolating was the only way to attempt controlling the spread until generalized vaccination became available. Testing also provided knowledge on transmission dynamics and variant detection, as well as anticipating surges in hospitalized cases. Testing was even more crucial in countries aiming for zero-COVID strategies. In such contexts, ensuring access to testing for the population is paramount. However, the distribution of tests can largely be heterogeneous in terms of time, space, but also population groups.

Recent studies analysed the link between deprivation and COVID-19 testing, incidence or morbidity-mortality at the national scale [5, 6]. These studies highlighted the combined role of deprivation and population density to increase COVID-19 burden. Deprivation was also associated with lower testing rates. However, these studies relied on deciles or quintiles of the national distribution of deprivation indices. These categories may not reflect accurately local disparities, due to different standards of living between regions (e.g. housing costs between Paris and Marseille, the two largest cities in France). Limits inherent to building indices may also bias results when applied at large scale, when specific situations are difficult to capture accurately. For example, well-off urban population may live without a car, and conversely rural populations across a wide range of socio-economic conditions likely own at least one car and live in a personal house. In addition, these studies only accounted for population density and did not adjust for access to healthcare, nor for the age structure of the population.

Our objective was therefore to analyse the relationship between socio-economic profile and SARS-CoV-2 testing and incidence rates during the different phases of the epidemic. We also aimed to support intervention allocation by the regional public health agency of the Provence Alpes Côtes d’Azur (PACA), a geographically heterogeneous region combining dense urban coastal regions and rural mountainous areas in South-Eastern France.

## Material and methods

### Study design

We conducted an ecological analysis of SARS-CoV-2/COVID-19 testing rate in the Provence-Alpes-Côte d’Azur (PACA) region in south-eastern France, at the highest spatial resolution available for aggregated epidemiological data in France: “regrouped islets for statistical information” (French acronym: IRIS used hereafter). IRIS correspond to contiguous geographical areas regrouping between 1000 and 5000 inhabitants. Municipalities (lowest local authority level) with a population <5000 inhabitants typically correspond to a single IRIS, while municipalities >5000 are divided into several IRIS. The PACA region counts 946 municipalities and 2,446 IRIS, corresponding to 5.04 million inhabitants.

### Study period

We analysed COVID-19 testing rate from the start of the second wave in the PACA region on 21 July 2020 to the regional upsurge in incidence corresponding to the onset of the omicron wave on 23 December 2021. We combined dates of implementation of public health measures and local incidence minima to account for the multiple distinct testing incentives or constraints faced by the population. We distinguished 10 epidemic periods (Figure S1, Table S1). The delimitations were: the two nationwide lockdowns starting dates (30 October 2020 and 24 March 2021) and first easing up dates (28 November 2020 and 3 May 2021); the end of Christmas holidays (4 January 21); the lowest regional incidence in June before delta-variant wave 4 (23 June 2021); the decree establishing a mandatory health pass for recreational and cultural events >50 participants (16 July 2021), which required a complete vaccination or a negative test result of less than three days, or evidence of a COVID-19 infection for >10 days and ≤6 months; end of convenience test gratuity for unvaccinated individuals (15 October 2021); the regional onset of the delta variant-associated fifth wave (8 November 2021).

### COVID-19 tests and cases data

COVID-19 tests and confirmed cases were available as 7-day cumulative counts aggregated by IRIS from the French National Public Health Agency (Santé Publique France) SI-DEP information system, which aggregates results of all SARS-CoV-2 RT-PCR and antigenic tests performed in France. People presenting for a test provided their home address systematically and a pre-processing algorithm mapped them to the corresponding IRIS.

### IRIS (spatial unit) selection

We excluded IRIS with ≤ 30 inhabitants because of incomplete covariate data due to non-disclosure of local income statistics when the number of inhabitants is insufficient to preserve anonymity. In addition, we excluded “activity” IRIS hosting >1000 workers during the day with twice as many workers as inhabitants, as well as “diverse” IRIS corresponding to low population areas (e.g. a protected natural area in the periphery of a city), because the resident population profile could be very different from the actual population frequenting and influencing the transmission. We also excluded IRIS where the average monthly number of tests exceeded three times the actual population over multiple periods, due to likely address errors in laboratories.

### IRIS (spatial unit) descriptive data

#### Sources

We obtained data describing the population of each IRIS from the French National Institute of Statistics and Economic Studies (INSEE). We used the national census database, which provides descriptive data on the population by IRIS and the equipment public database, which provides an exhaustive list of equipment located in each unit, with geographical coordinates (Table S2).

#### Socio-demographic variables

We characterized the population inhabiting each IRIS using the following variables: (i) percentage of the population ≥15 years old in each social and professional categories (8-category job classification: agriculture, business owners/independent, white-collar, intermediate, employees, blue-collar, pensioned/retired, unemployed/other-including students); (ii) percentage of total IRIS population of foreign origin; (iii) percentage of immigrants in total IRIS population; (iv) four variables used to calculate the French deprivation index (percentage of high-school graduates in population ≥15 years old not studying; percentage of unemployed in the active 15-64 years old population; percentage holding a blue-collar job in the active 15-64 years old population; median income) [7]; (v) proportion of overcrowded main residences; (vi) European deprivation index (EDI), which combines ten census-based variables aggregated at the IRIS-level, and deprivation variables at the individual level (proportion of individuals of foreign nationality, of households without a car, of individuals employed as managers or intermediate professionals, of single-parent families, of households with at least two individuals, of non-owner-occupied households, of unemployed individuals, of individuals without post-secondary school education, of overcrowded dwellings, and of non-married individuals) [8]; (vii) population density; and (viii) percentage of inhabitants belonging to 4 age groups: <18, 18-39, 40-64, >65.

#### Access to healthcare variables

We separately considered the general access to healthcare and the specific access to SAR-CoV-2 tests.

We characterized the general access to healthcare at IRIS level as the number of primary healthcare practitioners (medical doctor (MD), nurse, physiotherapist…) present in the IRIS. We also used localized potential accessibility (LPA), an indicator defined at municipal level corresponding to the number of MD consultations available per year for each person based on their residence (Table S2). This composite indicator takes into account the number of MDs relative to the population in the corresponding catchment area, population expected needs and the travel time to the nearest MD.

We considered the specific access to SAR-CoV-2 tests following two main options available for general population testing: medical laboratories, which conducted RT-PCR-based tests throughout all periods; and pharmacies, which deployed antigenic testing from period 3 onwards. We considered distance from a IRIS to the nearest facility and the number of facilities in the IRIS. Preliminary analysis showed a strong correlation between distance to pharmacies and distance to laboratories (Spearman correlation coefficient=0.6); and the minimal distance to either facility corresponded to the distance to a pharmacy (Spearman correlation coefficient=1) (Figure S2). Numbers of pharmacies and laboratories were also correlated (Spearman correlation coefficient=0.4).

Exploratory analysis of all variables by Spearman correlation confirmed by principal component analysis (PCA) indicated strong positive correlations between general access to healthcare and specific access to testing: a negative correlation between distance to testing facilities and LPA, and strong correlations between the numbers of primary healthcare practitioners and laboratories or pharmacies in IRIS. There was only limited correlation between number of equipment and LPA or distances to testing facilities (Figure S2). As a result, we used LPA and number of primary healthcare practitioners in our main analyses, and replaced LPA with distance to pharmacy in the sensitivity analysis.

### Statistical analysis

Statistical analyses were performed using R software (version 4.0.5., R Core Team 2020. R Foundation for Statistical Computing, Vienna, Austria) and packages {mgcv}, {factominer} and {sf}.

#### Socio-demographic and age profiles

In order to evaluate socio-demographic characteristics of, we grouped IRIS into one age- and one socio-demographic profile using an unsupervised clustering method based on PCA followed by hierarchical clustering on principal components (HCPC) [9, 10]. We defined socio-demographic profile using all socio-demographic variables except age variables, and we generated age profiles separately using the 4 age variables. We studied direct effects of EDI, proportion of inhabitants older than 65 years and population density in a sensitivity analysis.

#### Variable selection

Variable selection in the multivariable model was done based on prior assumptions using a directed acyclic graph (Dagitty v3.0, Figure S3) [11]. For testing rate, we considered that two main factors could explain the testing rate in a IRIS in the different periods: first, characteristics of general accessibility to tests in the IRIS, due to its geographical location and pre-epidemic access to healthcare and second, elements related to individual testing behaviour of IRIS population, such as age, presence of a retirement home, and socio-demographic characteristics.

#### Statistical model

We used a generalized additive multilevel model (GAMM, [12]) with a random-effect at municipality level to account for similarities in IRIS of the same municipality and for the LPA variable definition available at municipality-level only. As appropriate for count variables, we used a negative binomial distribution to take into account overdispersion, with a log link and included IRIS census-defined log population as an offset. We also included a Gaussian kriging smoother based on the geographical coordinates of each IRIS to account for spatial autocorrelation. Continuous variables were first tested without linear assumption (as splines) in univariate analysis. Sensitivity analyses were conducted by substituting distance to the nearest pharmacy to LPA, as these indicators were too correlated for all to be included in the same model (see supplement). In a second sensitivity analysis, we adjusted for population density and proportion of population above 65 years as spline individual predictors (not requiring linear approximation), and studied the effect of the social deprivation directly including EDI as a linear predictor or as a spline.

### Ethics

Access to information was controlled and SI-DEP data were obtained in accordance with privacy laws (General Data Protection Regulation [EU] 2016/679). Clearance was obtained through a specific convention (number 22DIRA41-0) between Aix-Marseille University and Santé Publique France, from the Aix-Marseille University Ethic committee (number 2022-10-20-006), and from the Aix-Marseille University Data Protection Officer (number 513087).

## Results

### Spatial unit selection

Out of the 2,446 IRIS in the Provence-Alpes-Cote d’Azur region, we analysed 2,306 after excluding 74 activity IRIS, 40 diverse IRIS, 25 residential IRIS with population <30 inhabitants, and one single unit corresponding to a rural municipality with 36 inhabitants with monthly test rates >3 times larger than the IRIS population (Figure 1).

**Figure 1:**
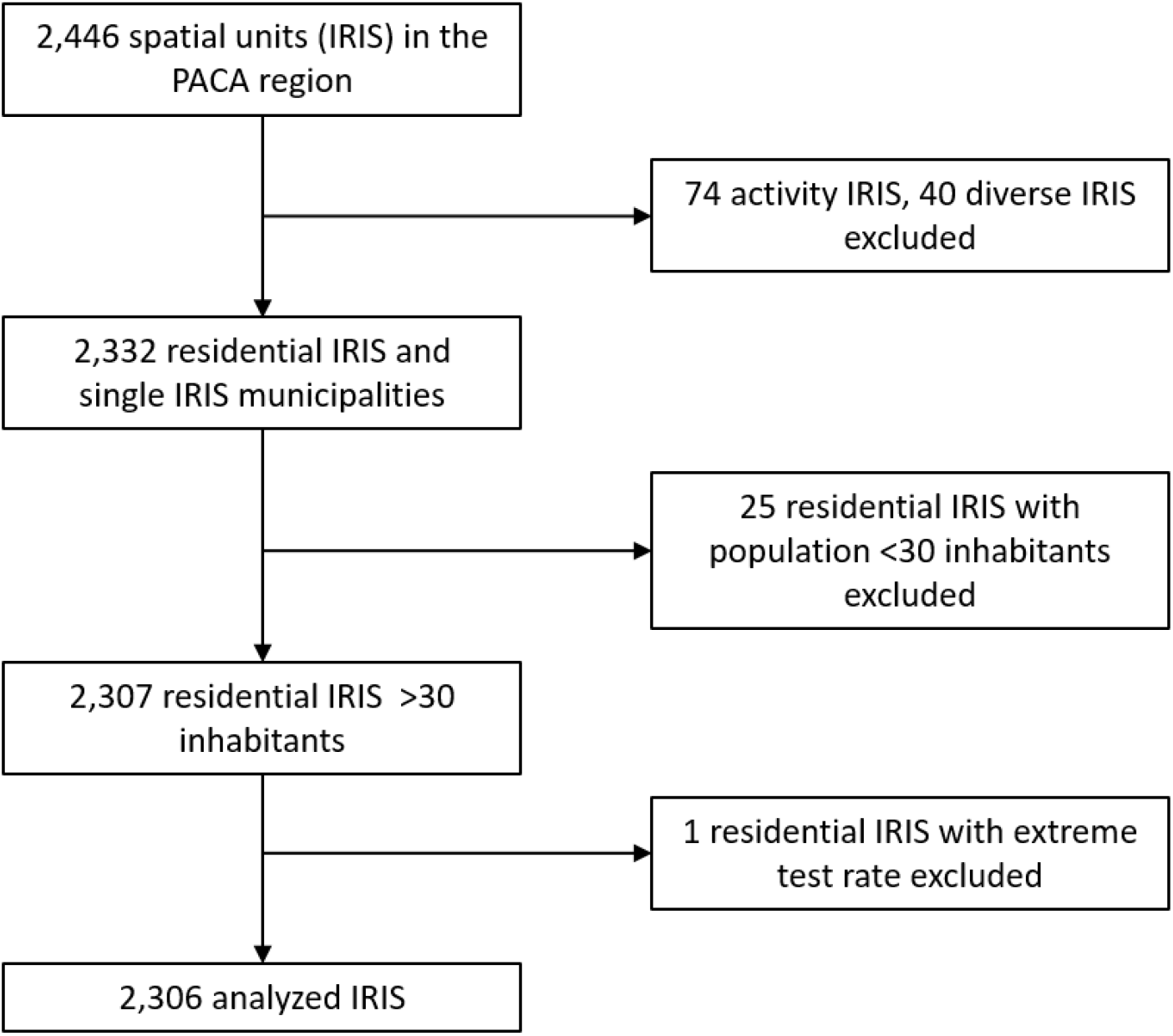
IRIS (spatial units) selection flow chart.

### IRIS (spatial unit) profiles

IRIS classification for socio-demographic variables identified 6 profiles ordered by increasing deprivation and exhibiting strong contrasts in terms of population density (Figure 2A, 2B). An intermediate-density profile corresponded to the lowest EDI, high income, high proportion of white collar and densities ranging from peri-urban to urban (profile 1, “privileged”). A very low-density profile included an important proportion of agriculture workers (profile 2, “remote”) and a second intermediate-density profile ranged from rural to peri-urban IRIS (profile 3, “intermediate”). A high-density urban profile corresponding to young adults, intermediate income with a high proportion of white collars (profile 4, “downtown”). Another high-density urban profile corresponded to areas with a high proportion of blue collar, lower income and intermediate density (profile 5, “deprived”). The third and last urban profile corresponded to very deprived urban areas with highest EDI, highest densities i.e. neighbourhoods of large social housing projects (profile 6, “very deprived”). IRIS profiles presented less heterogeneity in terms of age, with only two profiles displaying higher (remote) or lower proportion (very deprived) of population ≥65 (Figure 2F). Access to healthcare variables by profiles generally reflected the urban vs rural accessibility issue, access to healthcare being highly variable for rural IRIS (“remote” + “intermediate”) profiles (Figure 2G).

**Figure 2:**
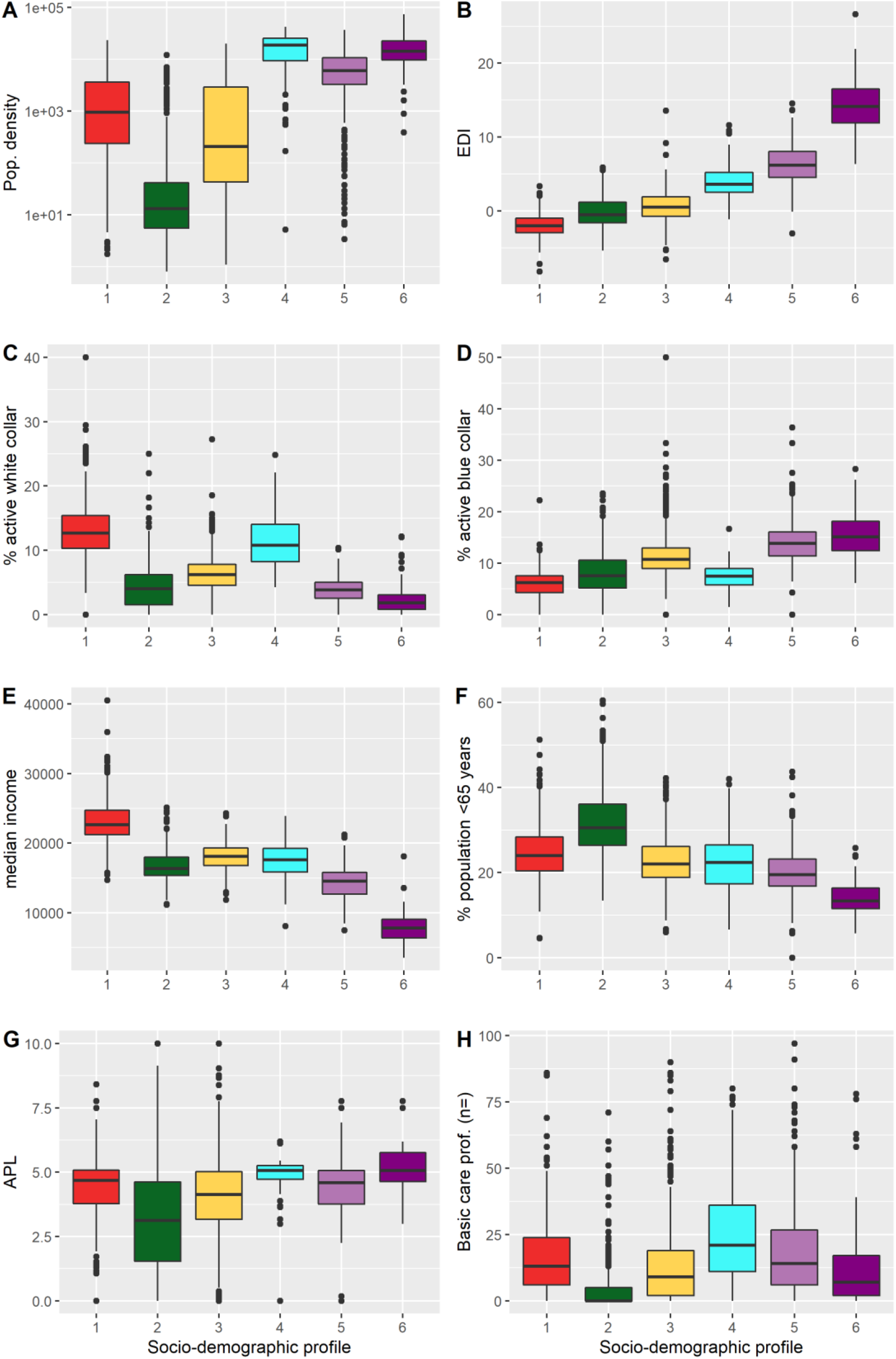
**Characteristics of the socio-demographic profiles of the spatial units (IRIS). (A) population density (log scale); (B) European Deprivation Index (EDI); (C) proportion of white collars among aged >15 years; (D) proportion of blue collar among the >15-year old population; (E) median income; (F) proportion of population aged >65 years; (G) localized potential accessibility (LPA) to healthcare indicator based on the average number of potential medical doctor visits available per inhabitant (defined at municipality level); and (H) number of primary healthcare professionals active in the census unit**. *Socio-demographic profiles: (1) “privileged” (red); (2) “remote” (green); (3) “intermediate” (yellow); (4) “downtown” (cyan); (5) “deprived” (light purple); (6) “very deprived” (dark purple). Corresponding histograms are presented in Figure S4*.

IRIS classification in age profiles identified 4 profiles. The three profiles “families”, “young adults”, and “elderly” were characterized respectively by a higher proportion of <18 years old; 18-39 years old, and ≥65 years old. The fourth profile, “balanced” exhibited similar proportions of inhabitants for each age category.

The geographical distribution of IRIS profiles matched expected patterns based on descriptive variables. “Remote” IRIS were mostly located in the mountainous areas of the region. “Privileged” IRIS mostly clustered in a vast area comprising and around Aix-en-Provence city, in the south and east of Marseille city, in along the coast between Marseille and Toulon city, along the coast between Cannes and Nice city. Lastly, “deprived” and “very deprived” IRIS concentrated in the northern part of Marseille city, or particular neighbourhoods of Toulon and Nice (Figure 3).

**Figure 3:**
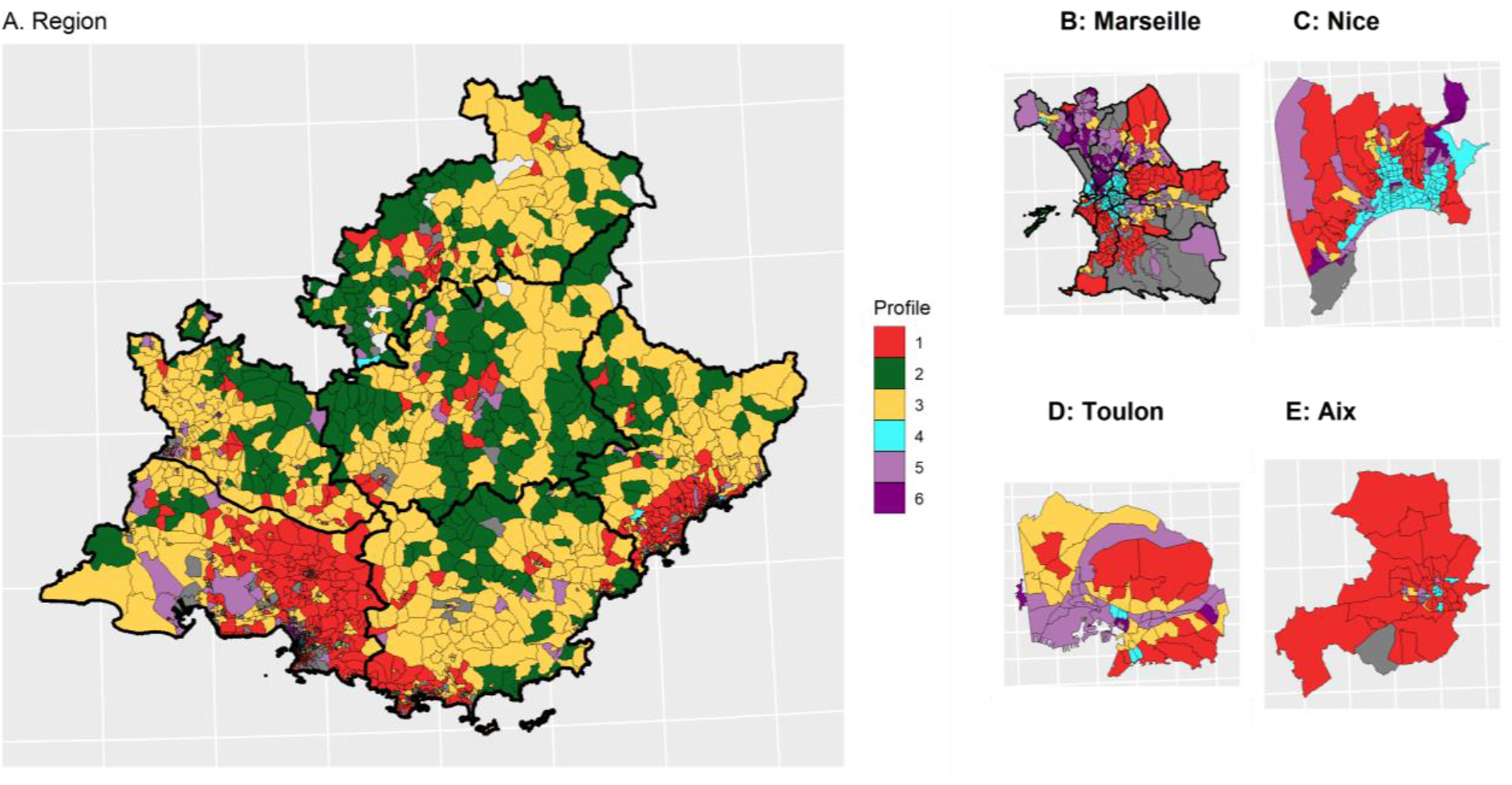
Spatial distribution of the socio-demographic profiles. (A) Provence Alpes Côte d’Azur (PACA) region; (B to E) main cities. *Socio-demographic profiles: (1) privileged (red); (2) remote (green); (3) intermediate (yellow); (4) downtown (cyan); (5) deprived (light purple); (6) very deprived (dark purple).*

### SARS-CoV-2/COVID-19 testing and incidence rates from July 2020 to December 2021 by socio-demographic profile

From July 2020 to December 2021, 5 peaks of SARS-CoV-2 testing rate were observed, in parallel with 4 waves of incidence (Figure 4). Testing peaks generally responded to increasing transmission periods, except for the 5^th^ testing rate peak that occurred during Christmas 2020. “Privileged” and “downtown” IRIS exhibited the highest testing rate overall, whereas the “very deprived” IRIS exhibited the lowest test rate, except during summer 2021 (after the health pass establishment) (Figure 4A). During that period, urban profiles (“downtown”, “deprived” and “very deprived”) displayed a general increase and “remote” or “intermediate” profiles showed the lowest testing rates.

**Figure 4:**
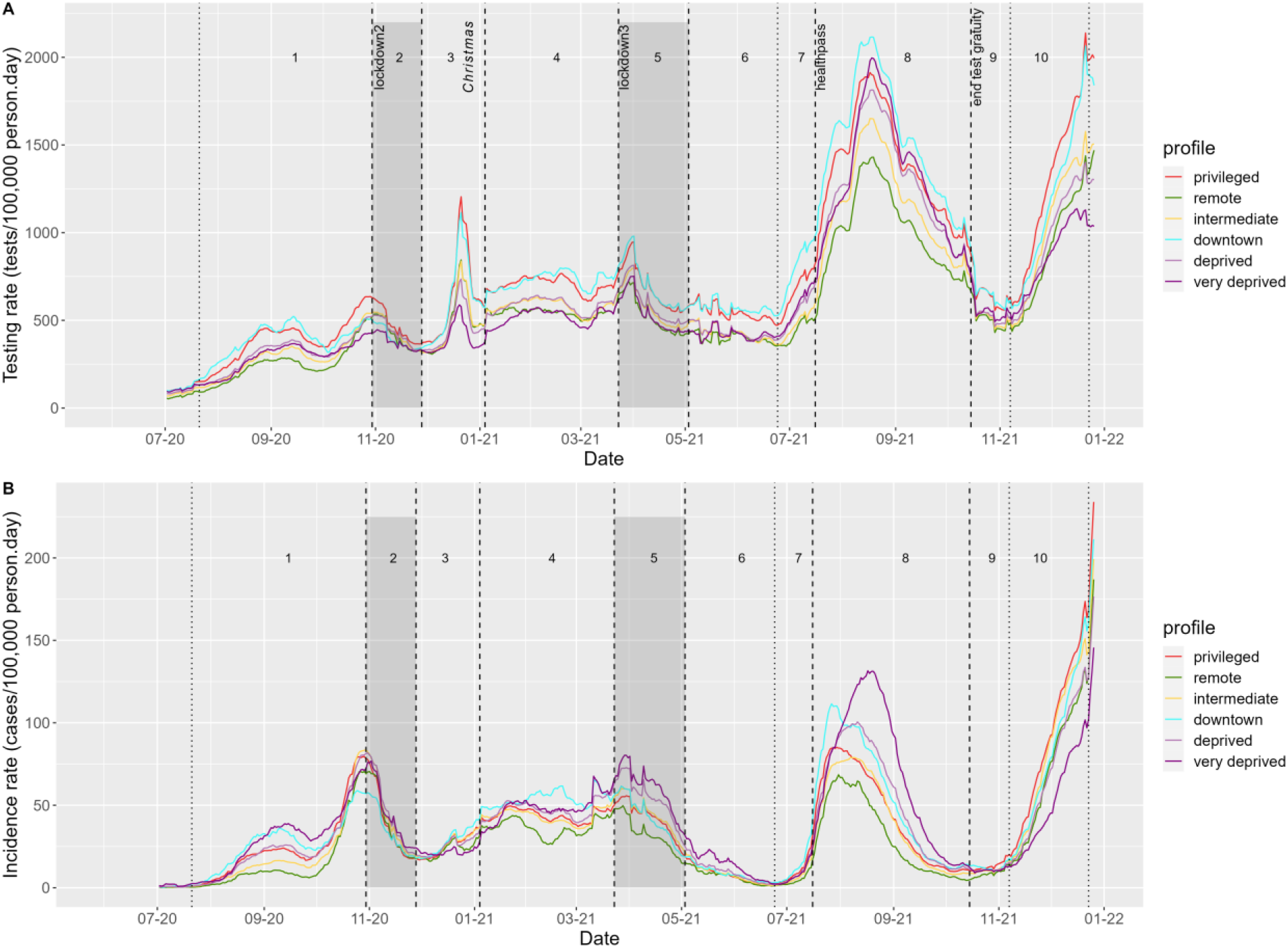
Evolution of daily SARS-CoV-2/COVID-19 (A) testing and (B) incidence rates, by sociodemographic profile in PACA region, from July 2020 to December 2021.

In comparison, COVID-19 incidence rates were highest in the “very deprived”, “downtown” and “privileged” units, despite contrasted testing rates (Figure 4B). The dynamics in these profiles were also different. In 2020, the “privileged” and “downtown” IRIS reached their maximum incidence rate one week before the October lockdown (period 1), while “very deprived” IRIS reached their peak during the week after the lockdown (period 2). Likewise, in July 2021 (period 8), “privileged” and “downtown” IRIS reached a maximum incidence rate at the end of July, compared to early August for “deprived” and mid-August for “very deprived” profiles, in a context of general high testing rates in these largely urban IRIS.

### Factors associated with COVID-19/SARS-CoV-2 testing rates during period 1

We analysed each period separately to identify factors associated with testing rate at IRIS level.

During period 1 (wave 2 rising), “privileged” IRIS showed a higher testing rate than all others and were chosen as a reference class. The adjusted testing rate ratio (aTRR) ranged from a 5% difference for “downtown” IRIS (aTRR=0.95, 95% confidence interval=[0.91-0.97]) to a 21% difference for “very deprived” IRIS (aTRR=0.79 [0.79-0.74]) (Table 1). The presence of elderly population also played a role, with an independent effect of the presence of a retirement home (aTRR=1.07 [1.04-1.09]) and of the elderly age profile (aTRR=1.11 [1.06 to 1.15], with “families” age profile as the reference class) (Table 1). LPA and number of primary healthcare professionals showed non-linear relationships with testing rate ratio (Table 1). For lower values of both variables, an increase was associate with a strong increase in aTRR. For LPA above 3 general practitioner consultations per inhabitant and per year, no additional effect on the aTRR was observed, while for IRIS with >10 primary healthcare practitioners, the increase of aTRR associated with supplementary practitioners was limited.

**Table 1:**
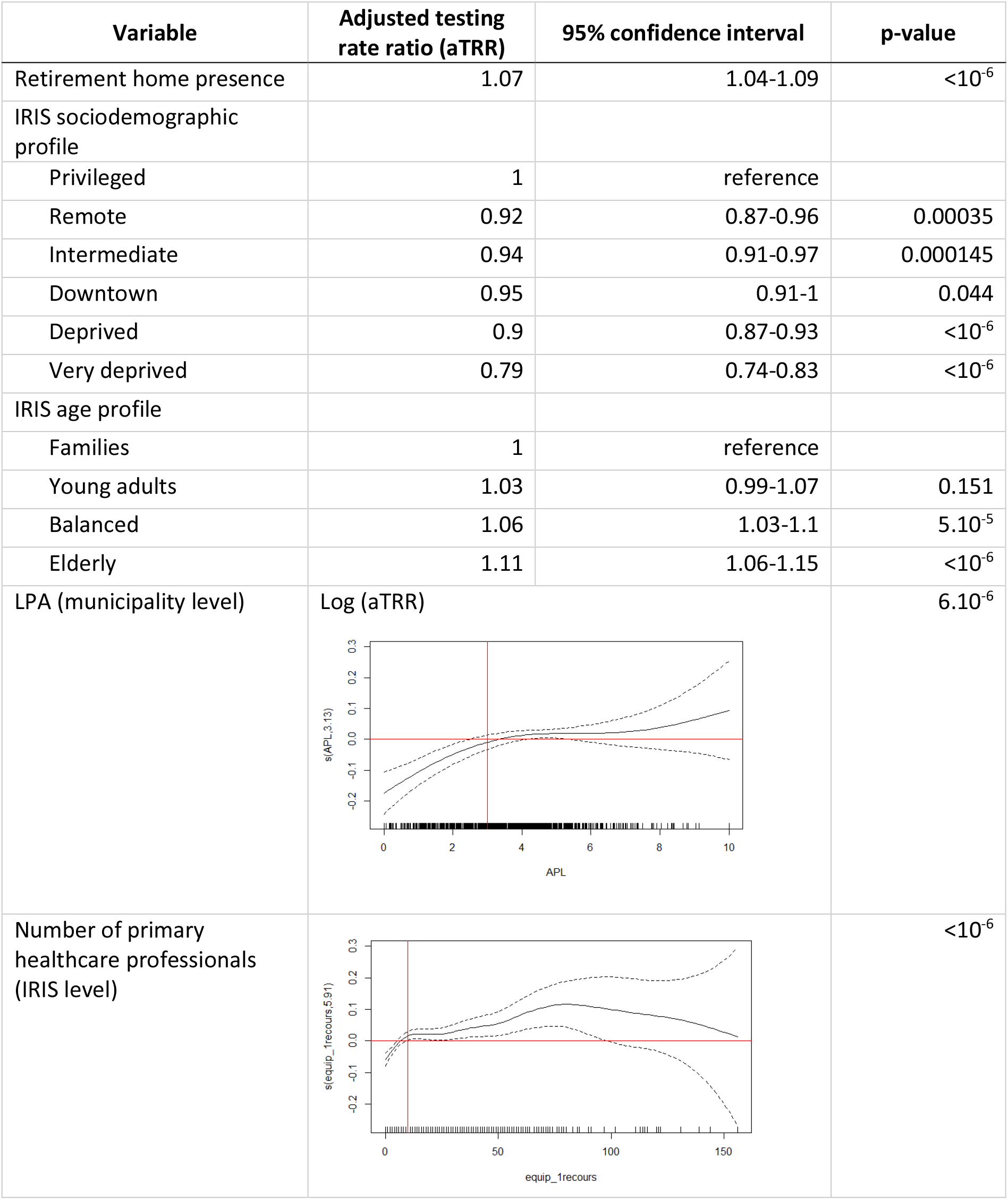
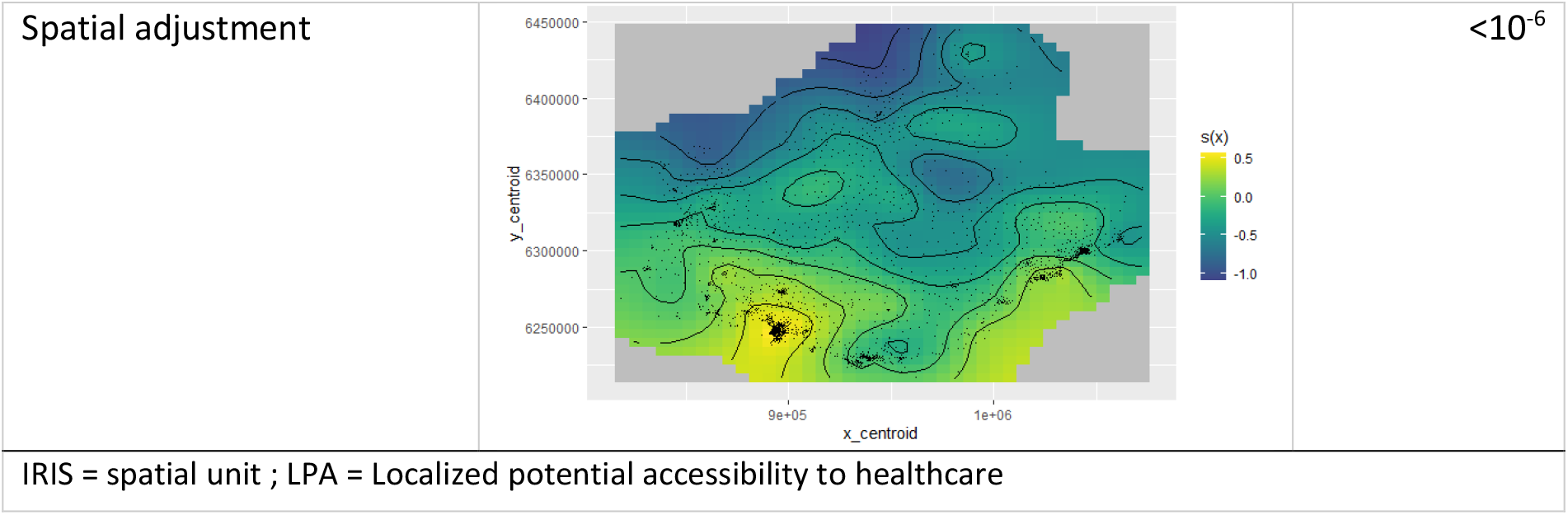
Adjusted factors associated with SARS-CoV-2/COVID-19 cumulative testing rate between 22 July and 29 October 2020: multivariate model results for period 1. *See Table S3 for univariate results*.

### Comparison of COVID-19/SARS-CoV-2 testing rates across socio-demographic profiles and periods

After adjusting for structural indicators of access to healthcare (LPA and number of primary healthcare practitioners) and spatial autocorrelation, “remote” and “intermediate” IRIS exhibited only limited gaps in SARS-CoV-2 testing rates compared to “privileged” IRIS (Figure 5A). “Intermediate” IRIS had higher testing rates compared to “remote” IRIS, with a parallel dynamic.

**Figure 5:**
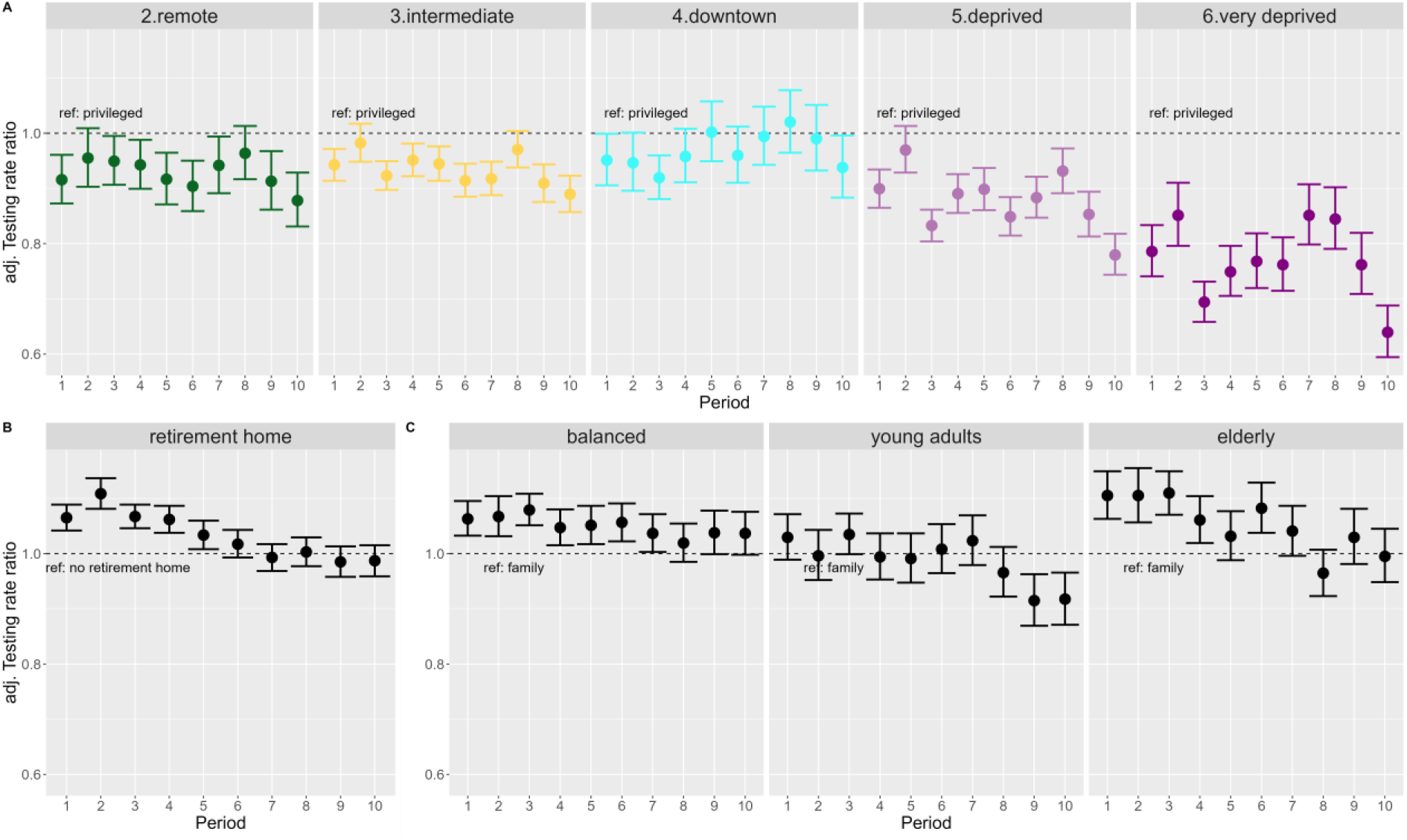
**Forest plots of adjusted testing rate ratios (aTRR) across all 10 periods: (A) changing patterns between socio-demographic profiles (reference class, “privileged” profile); (B) decreasing effect of retirement homes over time; and (C) changing patterns according to age profiles (reference class, “family” profile).**

Likewise, “downtown” IRIS did not have significantly lower aTRR except during period 3 and 10, corresponding to Christmas 2020 and 2021 periods (Figure 5A). On the other hand, aTRR were consistently lower for “deprived” IRIS (except during lockdown period 2) and for “very deprived” Sus (during all periods). The lowest differences (approximately 15%) between “privileged” and “very deprived” profiles were observed for periods 7 and 8 after health pass implementation. But in spite of massively available tests, periods 9 and 10 exhibited a sharp drop of testing in “deprived” and “very deprived” as compared to the “privileged” IRIS (Figure 5A).

The effect of the presence of a retirement home waned over the different periods (Figure 5B). The effect of age profile also changed gradually from early periods: elderly profile IRIS had higher testing than the family IRIS reference during early periods, and young adults profile IRIS had lower testing rate in the last two periods (Figure 5C).

The effect of LPA remained similar across periods, reaching a plateau around 2.5-3 available consults per inhabitant and per year (Figure S5). The effects associated to the number of primary healthcare professionals were more heterogeneous, with strongly non-linear shapes during the three “rising wave” periods 1, 7, 10 (Figure S6).

We conducted a sensitivity analysis replacing municipality-level LPA by IRIS-level distance to nearest pharmacy, which did not change the results (Figure S7). The direct analysis of EDI indicated that increasing deprivation was consistently associated with lower testing rates, but that this relationship was not linear for all periods. It also confirmed the periods of highest disparity between privileged and deprived profiles (Figures S8 and S9).

## Discussion

This geo-epidemiological study analysed factors associated with SARS-CoV-2 testing rates at the finest spatial scale available in the south-eastern French region across 10 periods spanning 18 months and corresponding to political measures (lockdowns, health pass), contextual events (Christmas) and epidemic waves. Our analysis showed strong contrasts in terms of socio-demographic profiles, access to healthcare, and population structure across the different periods analysed. It suggests that individuals living in “privileged” and “downtown” spatial units (IRIS) of the main regional cities sustained high testing rates. In contrast, individuals living in “remote” and “intermediate” IRIS displayed stable, slightly lower testing levels, after taking into account their more limited access to healthcare.

Looking away from large metropolitan areas, “remote” and “intermediate” IRIS presented parallel dynamics. While they exhibited only marginal differences in terms of income or EDI, and largely corresponded to rural areas, “intermediate” IRIS extended from rural to suburban areas with a higher population density and a better access to care (LPA and basic care professionals), whereas “remote” IRIS corresponded to smaller villages with low density, aging population, far from health services.

IRIS in “deprived” and “very deprived” profiles exhibited lower testing rates compared to “privileged” IRIS. Contextual testing periods (Christmas, periods 3 and 10) led to increasingly large testing gaps. Requirement of a health pass to access specific activities led to drastic testing increases in urban populations. However, the following period ending convenience test gratuity for unvaccinated asymptomatic individuals was associated with a dramatic drop in testing rates, mainly within “deprived” and “very deprived” areas, aggravating the underestimation of incidence rates.

Indeed, our results underline how testing disparities could affect the local monitoring of the epidemic: the highest incidence rates were recorded in “deprived”, “very deprived” and “privileged” profiles, however the epidemic situation could only be interpreted in the light of the much larger underestimation of cases in IRIS with higher levels of deprivation.

We studied drivers of SARS-CoV-2 testing at the finest spatial scale available in France, IRIS (or “IRIS” in French). We benefited from a wealth of contextual data provided by the French national census. Combining multiple data sources to characterize IRIS beyond population density, we could adjust for the general access to healthcare using different variables, and for the presence of elderly populations most likely to receive tests in the first periods before generalized vaccination. Presence of elderly populations was associated with a specific risk increase until the vaccination campaign reached sufficient coverage (period 5, ending in April 2021).

Access to healthcare is difficult to estimate at the IRIS level. Using a municipality-level localized potential accessibility (LPA) indicator may overestimate access in “very deprived” areas of metropolitan cities, where gaps in public transportation may isolate specific populations/neighbourhoods. On the other hand, many rural areas usually rely on the primary healthcare practitioners of the nearest town. Our strategy was thus to combine a distance-driven indicator (LPA or distance to pharmacy) and a presence-driven indicator (number of primary healthcare practitioners). This approach also allowed us to differentiate “remote” IRIS and “intermediate” IRIS, the latter showing a better access to care.

The IRIS-level vaccine coverage data was not available for our study, which precluded the analysis of factors associated with SARS-CoV-2 incidence rate after the onset of the vaccination campaign during the first quarter 2021. As a result, we could only show distinct incidence dynamics according to the IRIS profile, without quantifying the contribution of respective factors. We hypothesize that higher testing rates, better isolation abilities in more spacious housing, and enhanced ability to work remotely could explain the earlier incidence peak in “privileged” versus “very deprived” IRIS during period 2 and 5. In period 8, the different dynamics for the august peak are probably linked to differential vaccine coverage matching deprivation.

Based on our ecological approach, results excluded all individual components involved in population health behaviours. The decision whether or not to get screened stems from a particular psychological mechanism whose theoretical models are numerous [13] and whose determinants are not only associated with social characteristics, even if they play a certain role [14]. The simple economic dimension cannot by itself account for the complexity of health behaviours in the multiple dimensions in which they are deployed [15]; other elements must be considered in order to better understand them [16]. For example, some international studies have shown the influence of socio-cultural factors on health behaviours, and argue for further exploration of these [17]. Identified barriers to COVID-19 testing thus also include low health literacy, low trust in the healthcare system, or stigma and consequences of testing positive [18].

Our analysis displays stronger contrasts between most deprived and privileged areas compared to the analysis conducted at national level in France during comparable periods or in Switzerland [5, 6]. It also highlights the important contribution of elderly testing until April 2021. Our regional scale analysis based on socio-demographic profiles rather than national quintiles (France) or deciles (Switzerland) of deprivation indices allowed a precise characterization of local specific aspects : indeed, in our study region >30% of IRIS belonged to the highest deprivation quintile defined at French national level, respectively only 10% to the lowest quintile (Figure S10).

## Conclusion

We initiated this study in support of the regional public health agency of the PACA region (South-Eastern France) during the fourth quarter of 2020 to document the main drivers and inequalities in testing rate in the region and to identify areas with structurally low testing rates. Specific interventions (community engagement, home-visits for testing and supporting isolation efforts…) targeted these areas specifically. The “very deprived” IRIS profile was included as a contextual indicator in addition to weekly epidemic trends to prioritize health mediation interventions deployed by the regional health agency from October 2020 to June 2022 [19]. This fine spatial scale local profiling (infra-neighbourhood) is now included for general health mediation intervention projects in the city of Marseille.

## Data Availability

Generated Statement: The data analyzed in this study is subject to the following licenses/restrictions: SARS-CoV-2 testing and positive case data are government public health agency data not available openly. The aggregated data are accessible to researchers upon reasonable request for data sharing to the corresponding author. Requests for data require approval by Sante Publique France.

## Author’s contributions

JL and JG designed the study with contributions of SR. LB and JL collected and analysed the data with support of PC, FF and SN. All authors interpreted the results. JL, SR, MKBD, and JG drafted the first version of the manuscript. All authors contributed to the manuscript and reviewed the final version.

