## Supplementary file for "Social deprivation and SARS-CoV-2 testing: a population-based analysis in a highly contrasted Southern France region"

### Supplementary figures and results

#### 1 Supplementary material and methods

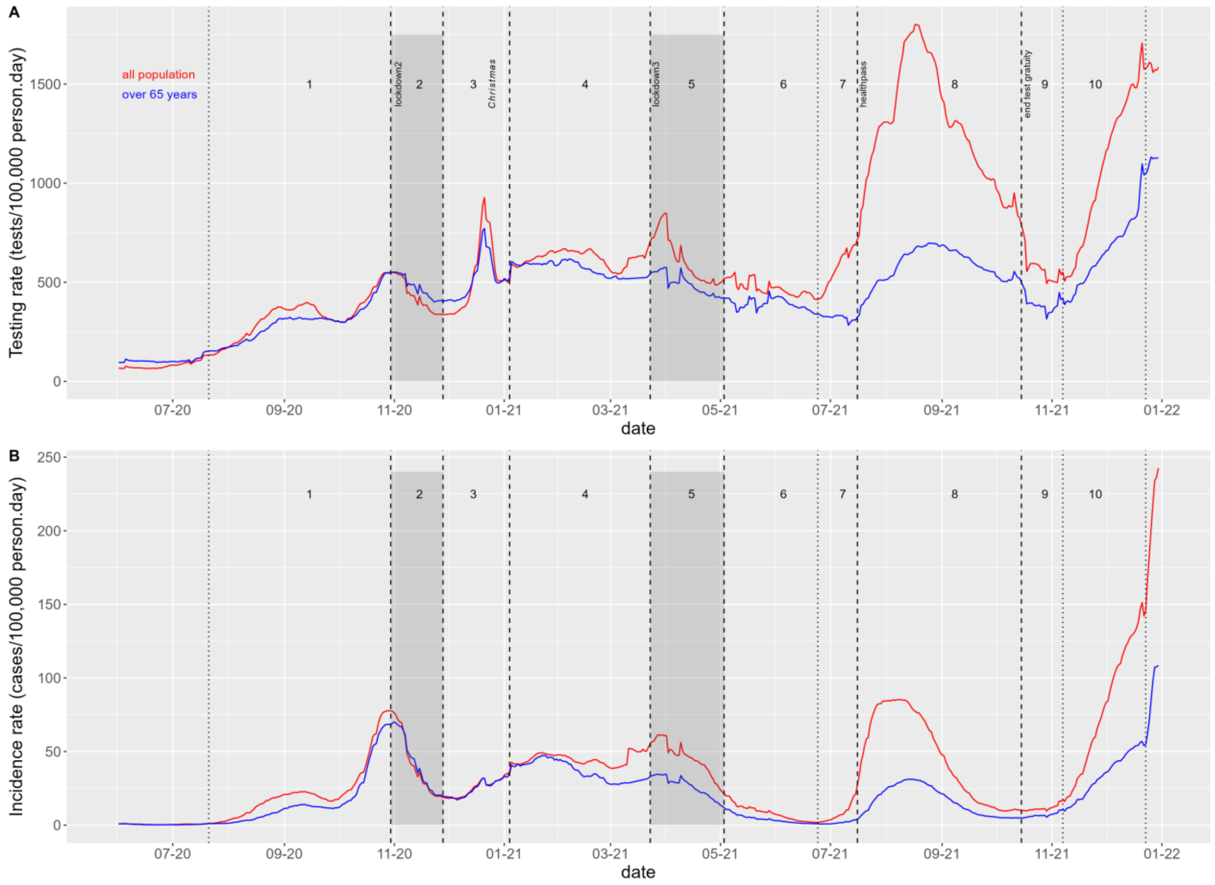

**Figure S1: Daily evolution of SARS-CoV-2/COVID-19 (A) testing and (B) incidence rates in the Provence Alpes Cote d'Azur (PACA) region in the general (red) and ≥65-years-old (blue) populations.**

**Table S1: Description of the ten epidemic periods of the COVID-19 epidemic in France considered for Provence Alpes Côte d'Azur (PACA) regional analysis between 22 July 2020 and 22 December 2021, i.e. from the onset of the second wave to the onset of omicron variant wave.**

| Period n° | Start and end dates | Starting Event | Total tests | Mean Testing rate (tests per 100,000 person.days) | Total cases | Mean Incidence rate (cases per 100,000 person.days) | Description |
|---|---|---|---|---|---|---|---|
| 1 | 22/07/2020 to 29/10/2020 | Incidence starts to increase again (wave 2) | 1,653,352 | 9,954 | 115,348 | 694.45 | Wave 2 rising |
| 2 | 30/10/2020 to 27/11/2020 | Lockdown date | 636,072 | 13,205 | 62,924 | 1306.32 | Lockdown 2 |
| 3 | 28/11/2020 to 03/01/2021 | 1st restrictions lifted | 968,744 | 15,763 | 46,956 | 764.05 | Christmas |
| 4 | 04/01/2021 to 23/03/2021 | End of Christmas holidays | 2,483,119 | 18,924 | 179,562 | 1368.42 | Wave 3 rising |
| 5 | 24/03/2021 to 02/05/2021 | Lockdown date | 1,240,047 | 18,664 | 92,356 | 1390.07 | Lockdown 3 |
| 6 | 03/05/2021 to 22/06/2021 | 1st restrictions lifted | 1,217,022 | 14,367 | 22,660 | 267.50 | Wave 3 falling |
| 7 | 23/06/2021 to 15/07/2021 | local incidence minimum (wave 4) | 646,405 | 16,920 | 8,384 | 219.46 | Wave 4 rising |
| 8 | 16/07/2021 to 14/10/2021 | Decree instituting mandatory healthpass | 5,828,291 | 38,560 | 214,360 | 1418.19 | Wave 4 healthpass |
| 9 | 15/10/2021 to 07/11/2021 | End of convenience testing gratuity for unvaccinated population | 698,027 | 17,510 | 14,103 | 353.78 | End of test gratuity |
| 10 | 08/11/2021 to 22/12/2021 | Onset of wave 5 (delta variant) | 2,440,882 | 32,656 | 181,676 | 2430.62 | Wave 5 Delta |

**Table S2: Spatial unit ("IRIS") covariate data sources**

| Database name/type | Date of publication | Source | URL |
|---|---|---|---|
| **Permanent equipment database 2020** | 12/07/2021 | INSEE | https://www.insee.fr/fr/statistiques/3568629?sommaire=3568656&q=bpe+2020 |
| **Census database** | 09/12/2020 | INSEE | https://www.insee.fr/fr/statistiques/4515565?sommaire=4516122&q=recensement+2017#consulter |
| **Available income database 2017** | 29/12/2020 | INSEE | https://www.insee.fr/fr/statistiques/4479212#consulter |
| **Housing database 2017** | 09/12/2020 | INSEE | https://www.insee.fr/fr/statistiques/4515532?sommaire=4516107&q=base+logement#dictionnaire |
| **European deprivation index (EDI)** | 2015 | MapInMed | https://anticipe.eu/plateformes/MAPinMED |
| **French deprivation index** | 01/04/2019 | INSERM | https://public.opendatasoft.com/explore/dataset/indice-de-defavorisation-sociale-fdep-par-iris/information/?flg=fr&q=croix&location=13,50.67628,3.13308&basemap=jawg.streets |
| **IRIS map (shapefile)** | | French Government | https://www.data.gouv.fr/en/datasets/decoupage-iris-combine-aux-limites-municipales-openstreetmap/ |
| **Localized potential access (LPA)** | 02/03/2020 | DREES | https://drees2-sgsocialgouv.opendatasoft.com/explore/dataset/530_l-accessibilite-potentielle-localisee-apl/information/ |

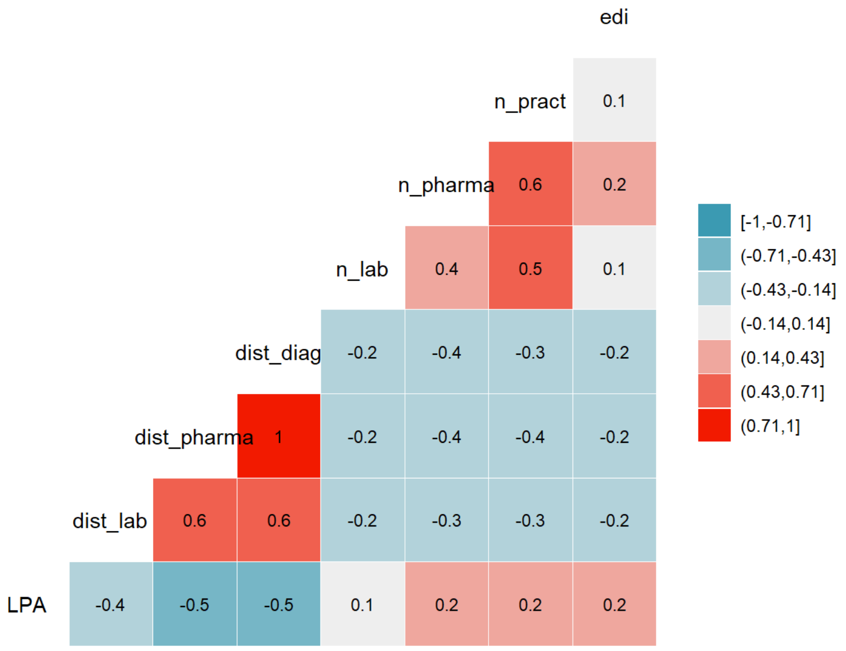

**Figure S2: Spearman cross correlation matrix between health access variables.** LPA= localized potential accessibility, i.e. number of general practitioner consults available per person and per year (calculated at municipal level). dist_lab = distance to the nearest laboratory performing biological exams. dist_pharma = distance to the nearest pharmacy. dist_diag = distance to the nearest equipment, combining laboratory and pharmacy. n_lab = number of laboratories in IRIS. n_pharma = number of pharmacies in IRIS. n_pract = number of primary healthcare practitioners in IRIS. edi = European deprivation index in IRIS (showing limited overall correlation with accessibility)

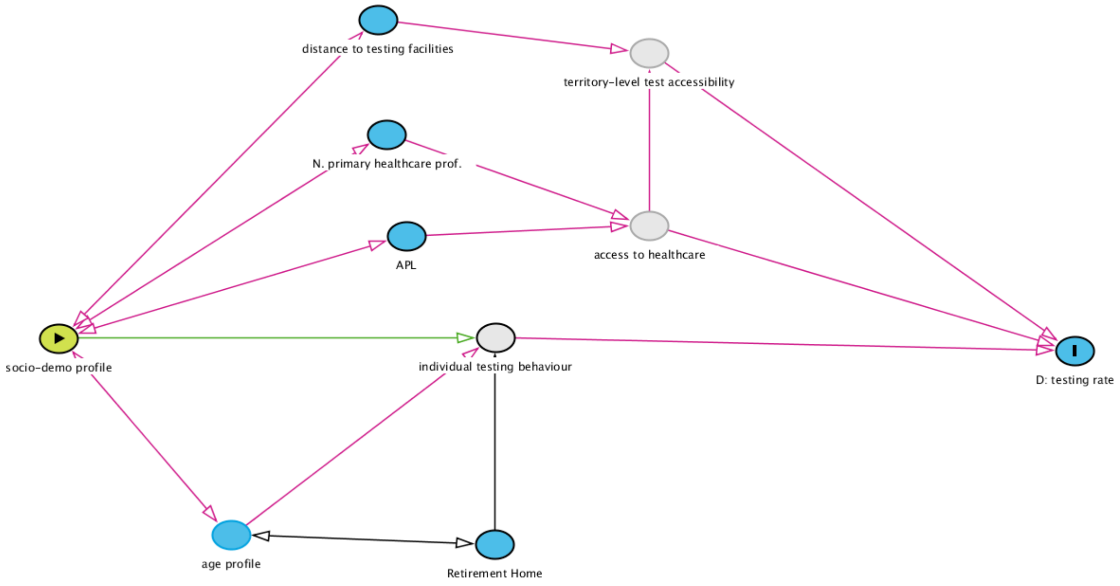

**Figure S3: Directed acyclic graph used to define the relationship between socio-demographic/deprivation profiles and SARS-CoV-2 testing rate in Provence Alpes Côte d'Azur region.**

#### 2 Supplementary results

##### 2.1 Regional distribution of IRIS descriptive socio-demographic and health access variables

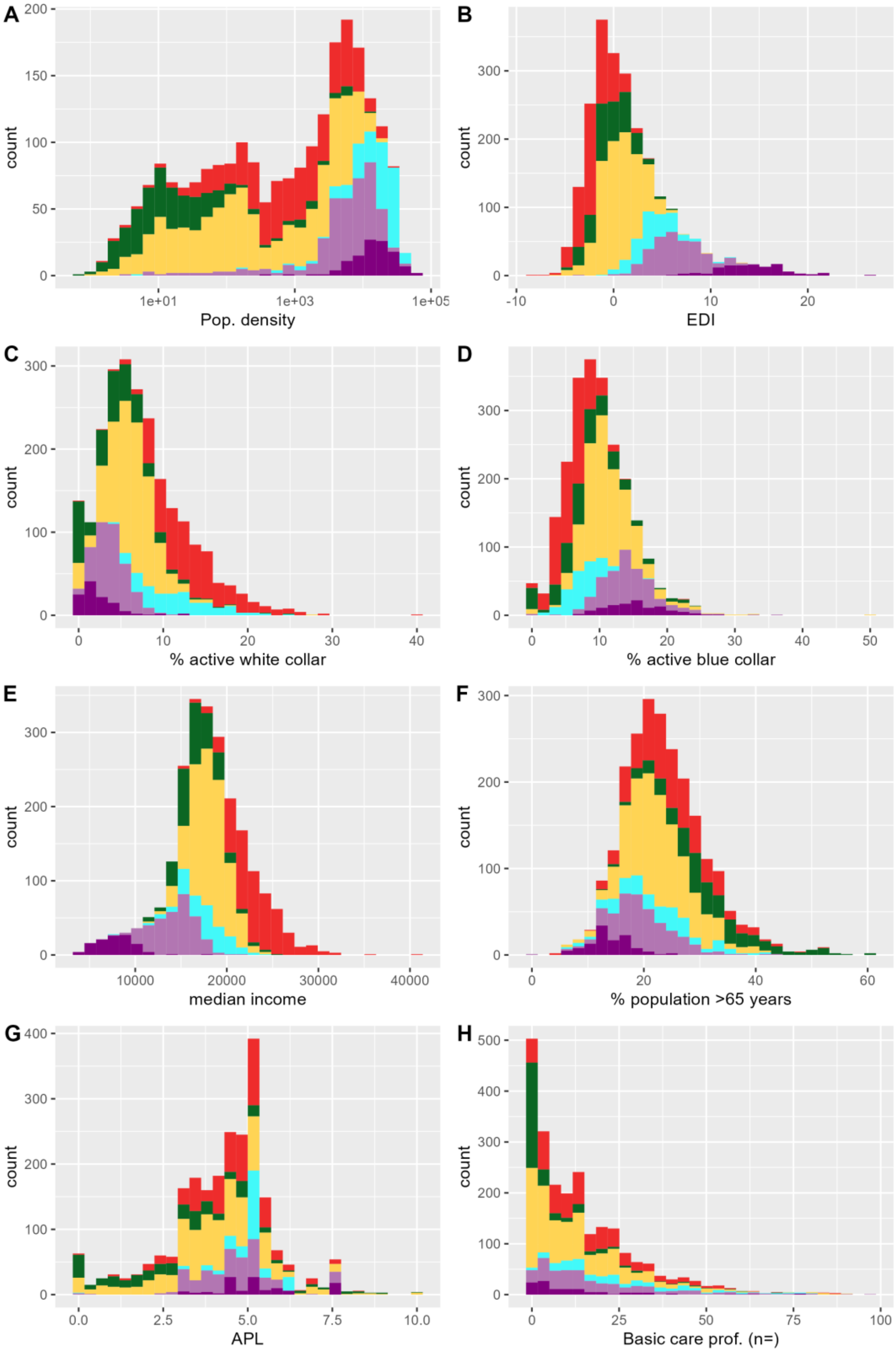

**Figure S4: Histograms of socio-demographic profile variables and health access variables, by profile.** Socio-demographic profiles: (1) “privileged” (red); (2) “remote” (green); (3) “intermediate” (yellow); (4) “downtown” (cyan); (5) “deprived” (light purple); (6) “very deprived” (dark purple).

#### 2.2 Univariate regression results for period 1

**Table S3: univariate results for period 1 (model including municipality-level random intercept and s(x,y) spatial autocorrelation adjustment)**

| Type | Variable | Category | TRR | 95% confidence interval | p-value | gcv | dev.expl |
|---|---|---|---|---|---|---|---|
| access to health-care | LPA | spline | 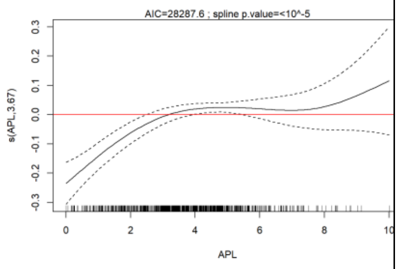  | | $<10^{-6}$ | 3778.0 | 74.3 |
| | n_pharma | 0 (ref) | 2.72 | (ref) | | 3784.9 | 74.4 |
| | n_pharma | n=1 | 1.03 | 1.01 1.06 | 0.009 | | |
| | n_pharma | n=2 | 1.05 | 1.02 1.08 | 0.0016 | | |
| | n_pharma | n>=3 | 1.14 | 1.09 1.20 | $<10^{-6}$ | | |
| | n_pract | spline | 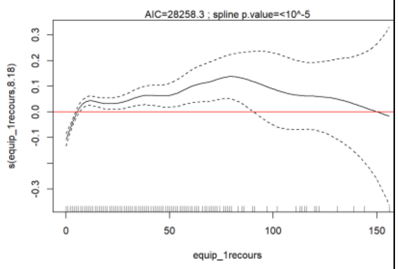  | | $<10^{-6}$ | 3791.5 | 74.6 |
| | s(dist_lab) | spline | 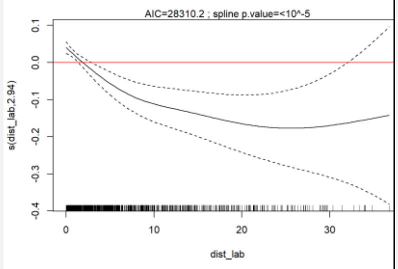  | | 0.000007 | 3778.9 | 73.9 |
| | s(dist_pharma) | spline | 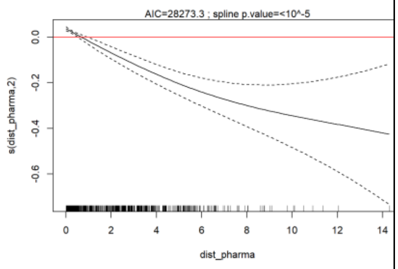  | | $<10^{-6}$ | 3763.5 | 74.1 |
| age | Age profile | families | 2.72 | ref | | 3832.3 | 75.7 |

| | | | | | | | |
|---|---|---|---|---|---|---|---|
| | | balanced | 1.12 | 1.09 1.15 | $<10^{-6}$ | | |
| | | retired | 1.18 | 1.14 1.22 | $<10^{-6}$ | | |
| | | young_adults | 1.08 | 1.04 1.12 | 0.000127 | | |
| | pop_aged_over65 | spline | 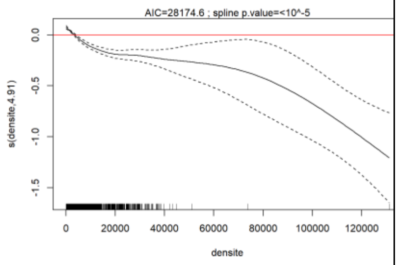  | | $<10^{-6}$ | 3818.4 | 75.4 |
| | retirement home | No | 2.72 | (ref) | | 3751.2 | 73.9 |
| | | yes | 1.10 | 1.08-1.13 | $<10^{-6}$ | | |
| s(edi) | s(edi) | spline | 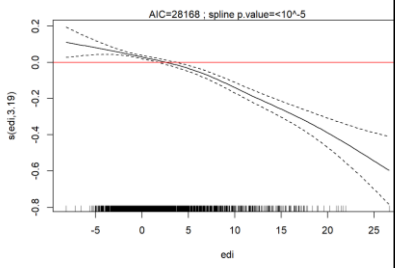  | | $<10^{-6}$ | 3890.6 | 76.8 |
| Socio-demo-graphic | National EDI quintile | Q1 (ref) | 2.72 | (ref) | | 3853.17 | 75.4 |
| | | Q2 | 0.98 | 0.94-1.02 | 0.339791 | | |
| | | Q3 | 0.95 | 0.91-1.00 | 0.030164 | | |
| | | Q4 | 0.95 | 0.91-0.99 | 0.022974 | | |
| | | Q5 | 0.89 | 0.85-0.92 | $<10^{-6}$ | | |
| | pop. density | spline | 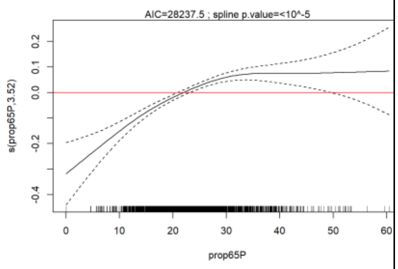  | | $<10^{-6}$ | 3867.0 | 76.8 |
| | SD profile | 1.privileged | 2.72 | (ref) | | 3850.5 | 75.9 |
| | | 2.remote | 0.89 | 0.85-0.93 | 0.000002 | | |
| | | 3.intermediate | 0.93 | 0.90-0.96 | 0.000003 | | |

| | | | | | |
|---|---|---|---|---|---|
| | | 4.downtown | 0.95 | 0.90-0.99 | 0.01598 5 |
| | | 5.deprived | 0.87 | 0.84-0.90 | $<10^{-6}$ |
| | | 6.very_deprived | 0.73 | 0.69-0.77 | $<10^{-6}$ |
| TRR, testing rate ratio; 95%-CI, 95%-confidence interval; LPA, Localized potential accessibility to healthcare; EDI, European deprivation index; SD, socio-demographic | | | | | |

#### 2.3 Multivariate, all periods, splines

##### *2.3.1 LPA splines*

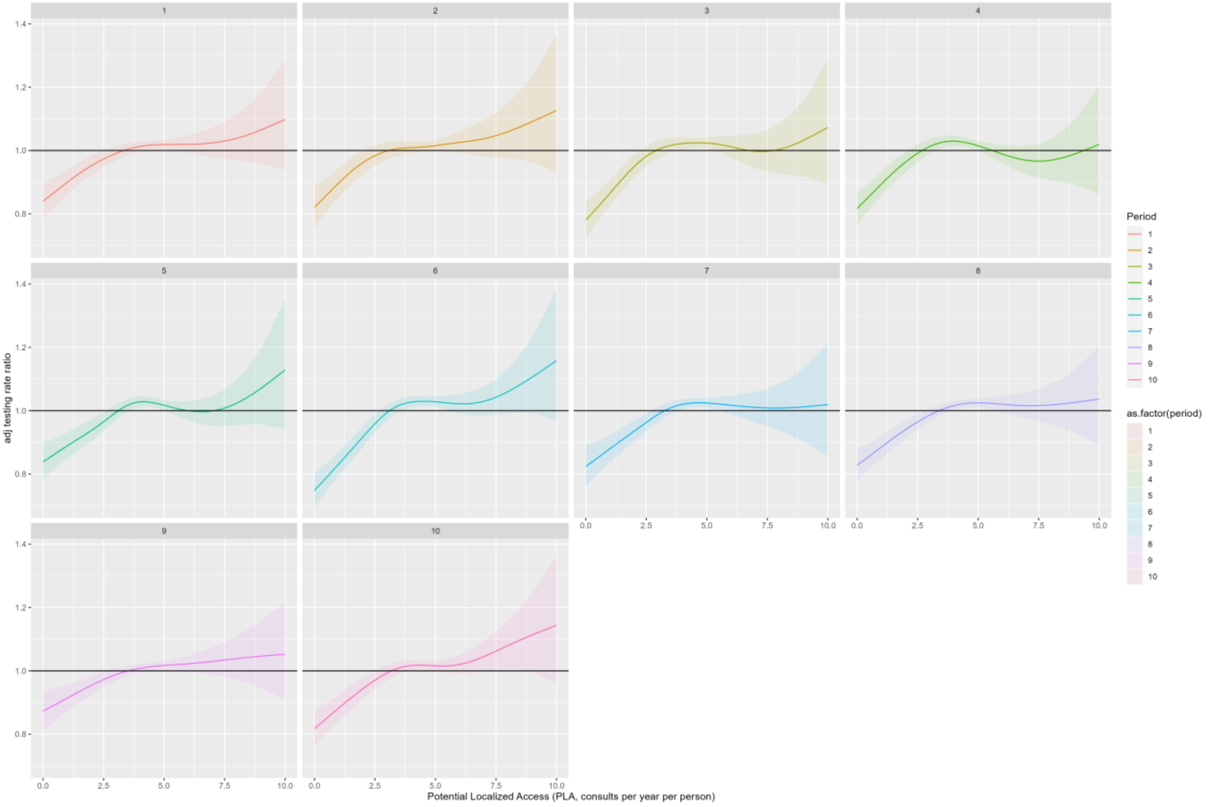

**Figure S5: Non-linear effect of localized potential accessibility to healthcare (LPA) for periods 1 to 10 (panels).**

##### 2.3.2 Number of basic healthcare profiessional slines

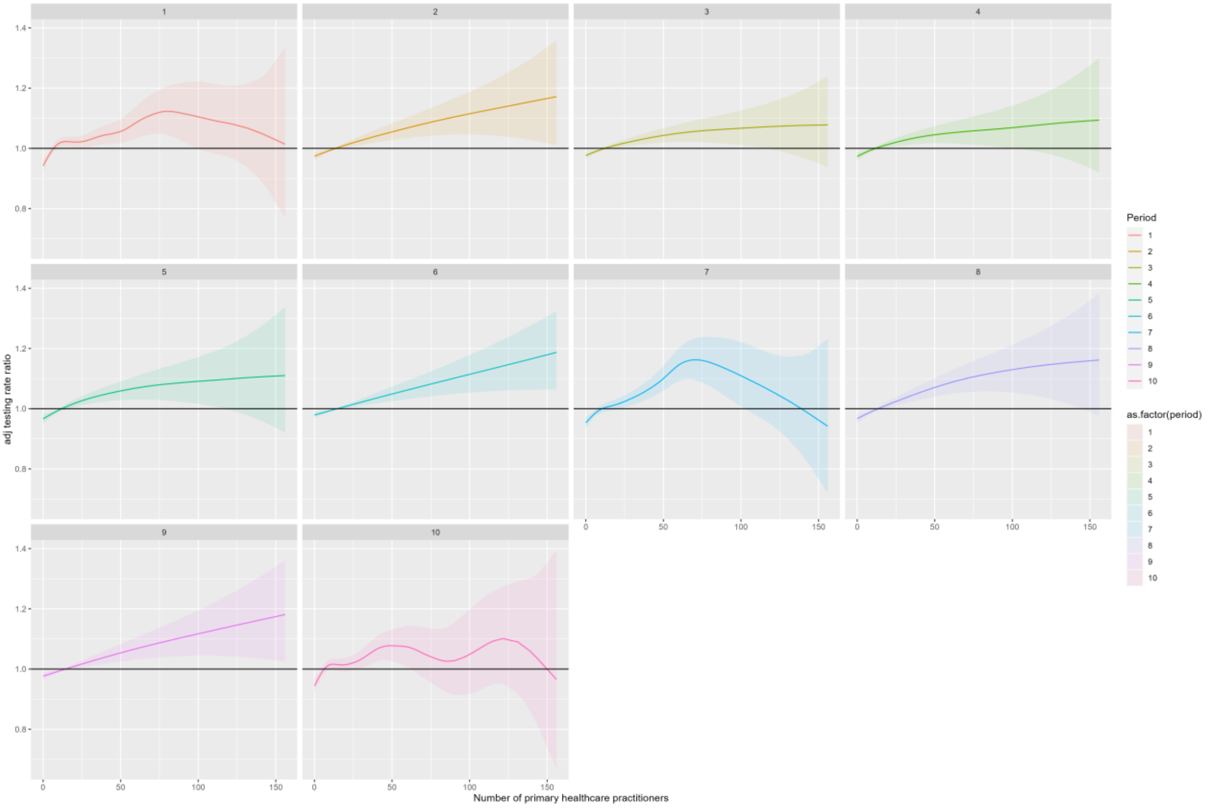

**Figure S6: Non-linear effect of the number of basic healthcare professionals for periods 1 to 10 (panels).**

#### 2.4 Multivariate, all periods, sensitivity analysis

##### *2.4.1 Replacing municipality-level LPA with distance to the nearest pharmacy*

Multivariate analysis performed replacing localized potential accessibility to healthcare (LPA) with distance to the nearest pharmacy, all other variables remaining identical.

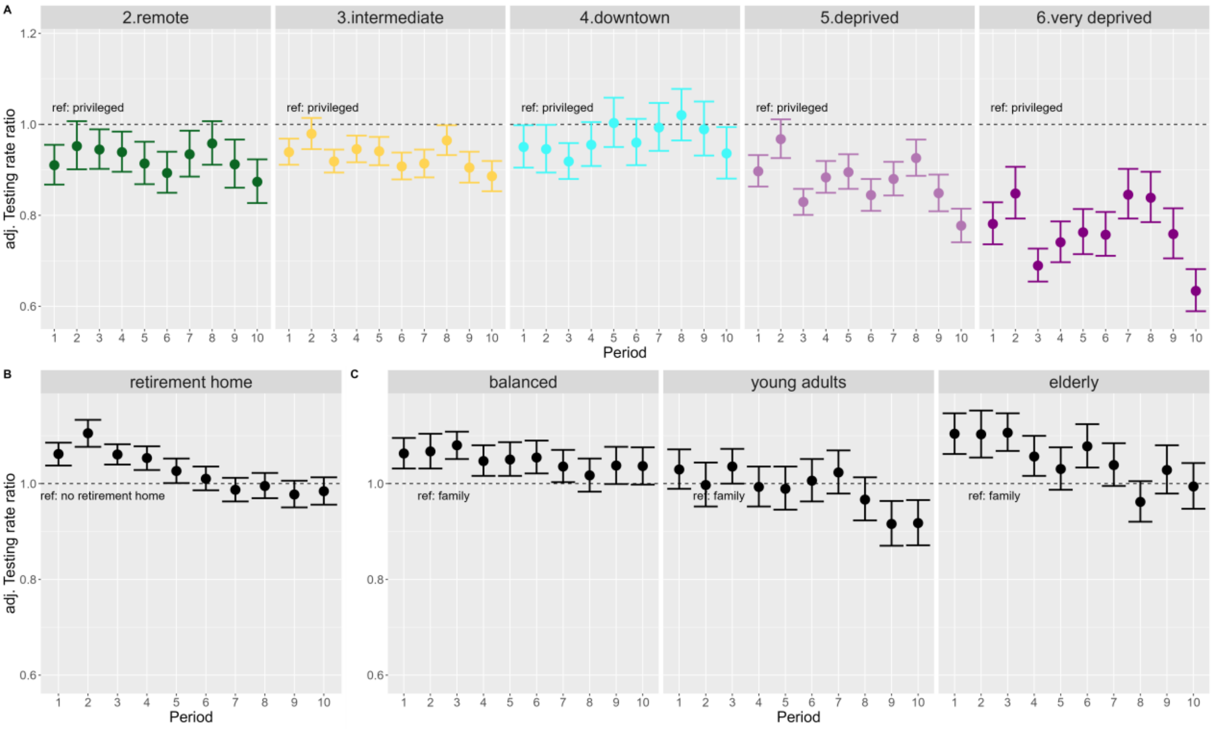

**Figure S7: Combined forest plots for all periods substituting distance to the nearest pharmacy to replace localized potential accessibility to healthcare (LPA).** One multivariate regression by period, all models combined for the 3 variables.

##### 2.4.2 Replacing socio-demographic profiles with EDI values

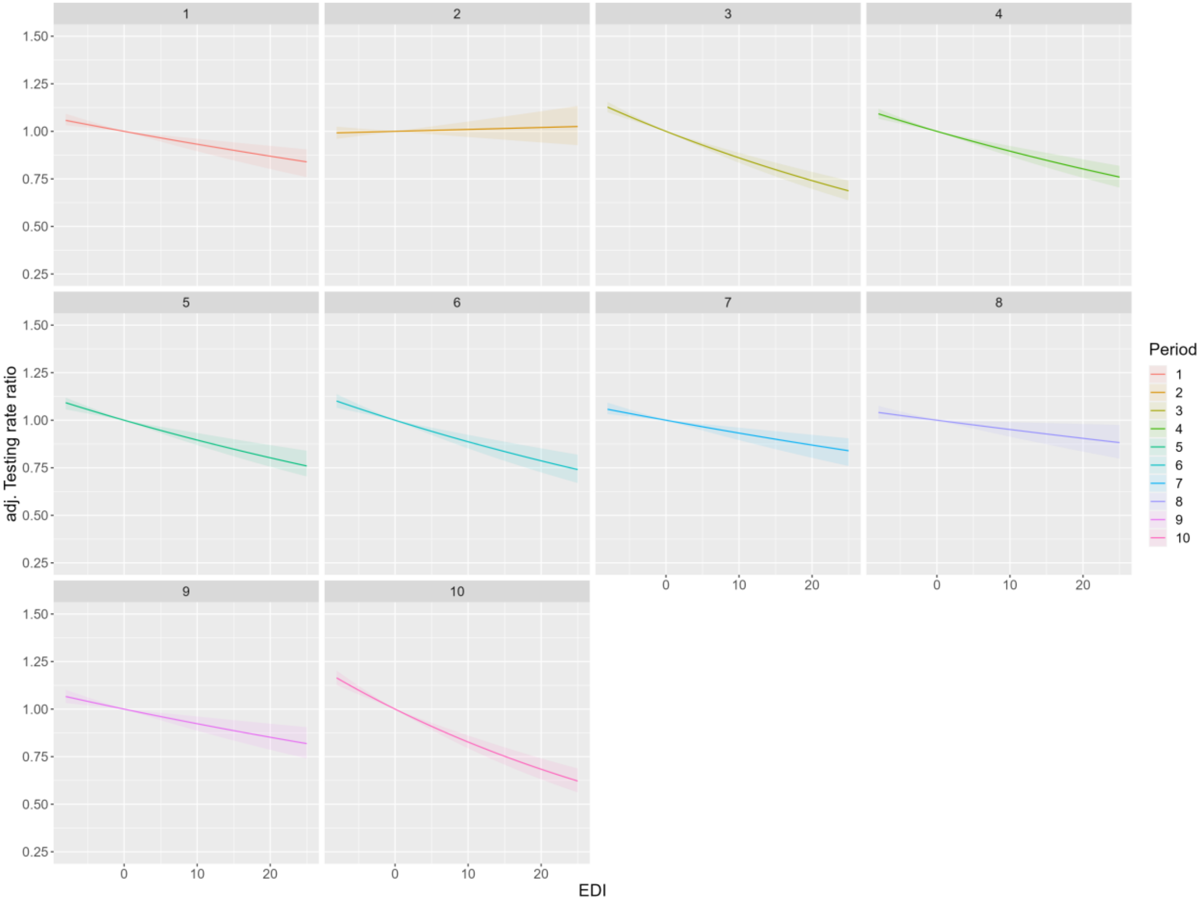

**Figure S8: Adjusted Testing Rate Ratio (aTRR) by European deprivation index (EDI) value for each period, assuming an exponential effect of EDI on testing rate ratio (TRR) (i.e. a linear effect of +1 unit of EDI on log(TRR)).** Multivariate negative binomial regression model including adjustments for localized potential accessibility to healthcare (LPA), number of primary care professionals, presence of a retirement home, proportion of the population above 65 years of age, population density, with a municipality-level random intercept and a Gaussian kriging smoother of the geographical coordinates of each IRIS.

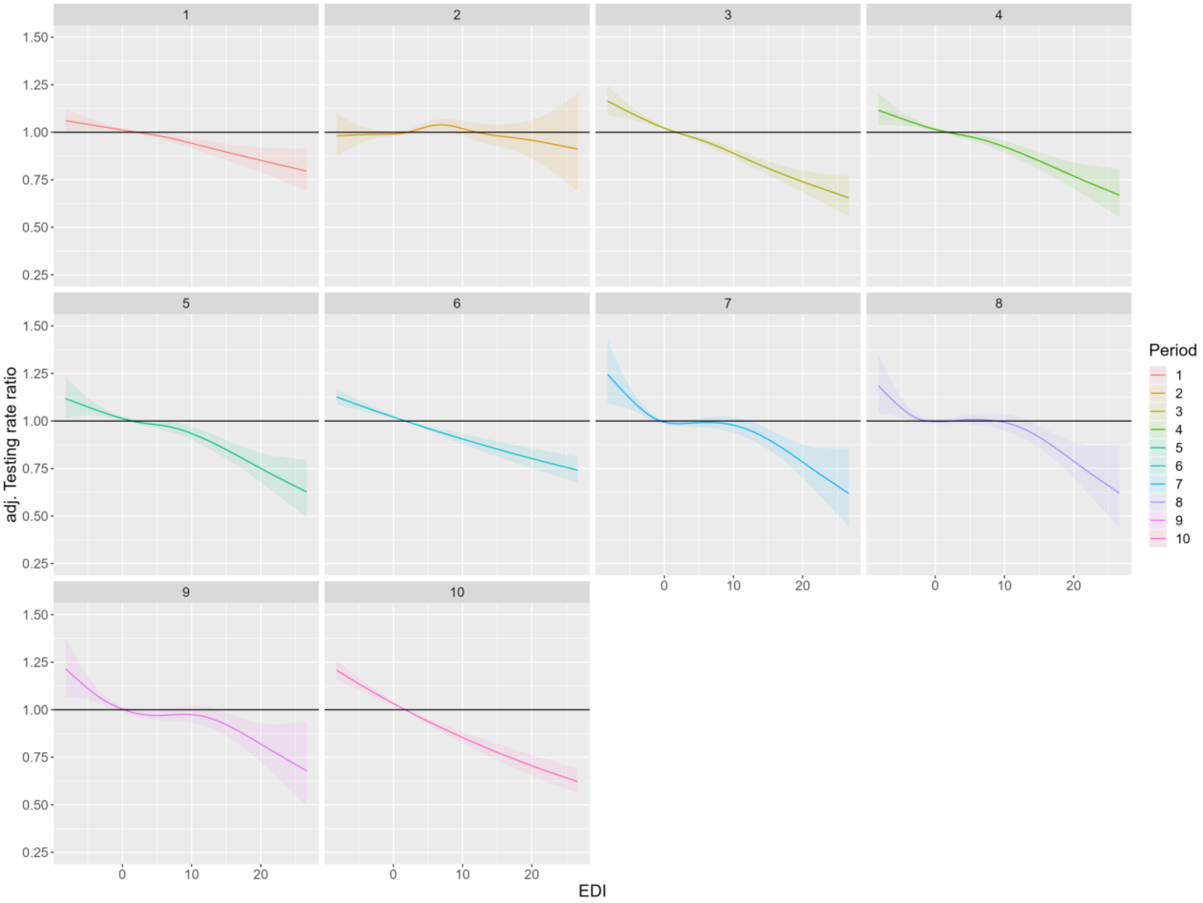

**Figure S9: adjusted Testing rate ratio (aTRR) by European deprivation index (EDI) value for each period, assuming a non-linear effect of EDI on testing rate ratio (TRR) (spline).** Multivariate negative binomial regression model including adjustments for localized potential accessibility to healthcare (LPA), number of primary care professionals, presence of a retirement home, proportion of the population above 65 years of age, population density, with a municipality-level random intercept and a Gaussian kriging smoother of the geographical coordinates of each IRIS.

#### 2.5 IRIS socio-demographic profile versus national deprivation index quintiles

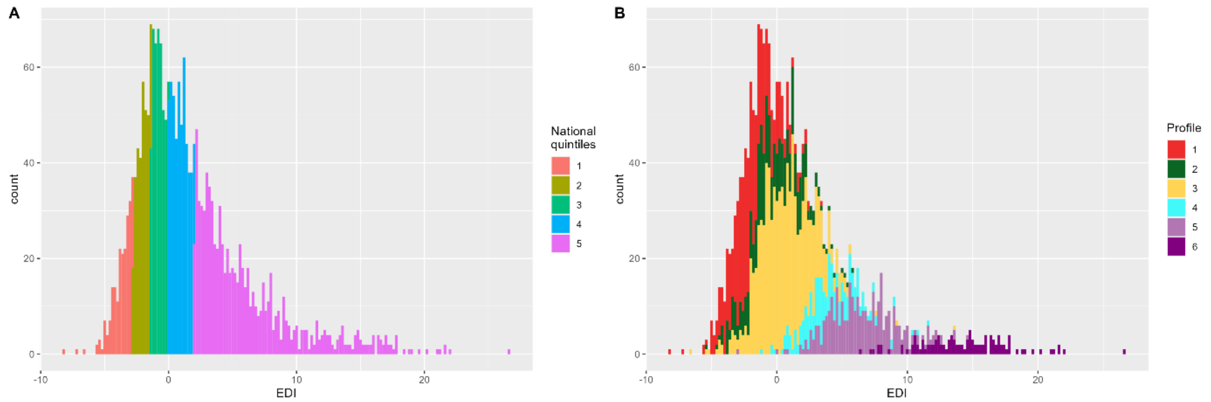

**Figure S10: Distribution of European Deprivation Index (EDI) value for 2306 analysed IRIS. (A) by national EDI quintile; (B) by socio-demographic profile as defined in this study.** Socio-demographic profiles: (1) “privileged” (red); (2) “remote” (green); (3) “intermediate” (yellow); (4) “downtown” (cyan); (5) “deprived” (light purple); (6) “very deprived” (dark purple).

The regional analysis of European Deprivation Index (EDI) using national quintiles would not bring sufficient precision. Indeed, of 2306 residential IRIS with >30 inhabitants, 36.51% (n=842) correspond to 5$^{th}$/highest national EDI quintile. Only 9.15% (n=211) are in the first quintile, 14.01% (n=323) in the second,17.91% (n=413) in the third, and 22.42% (n=517) in the fourth (Figure S4A). The fifth quintile actually covers 4 different sociodemographic profiles in the region, corresponding to very contrasted features: “very deprived” (urban, very dense); “deprived” (urban corresponding to both neighbourhoods in large metropolitan areas and smaller industrial towns for example); “downtown” (very dense, urban, with young adults and a significant proportion of white-collar workers); “intermediate” (suburban to rural areas) (Figure S4B).
